# Epidemiological pattern of births from the largest surveillance database of live births in Brazil “SINASC” before and during the COVID-19 pandemic in the Brazilian Amazon

**DOI:** 10.1101/2022.09.02.22279455

**Authors:** Daniele Melo Sardinha, Andréa Maria da Silva Luz, Kátia Raquel Almeida Carneiro, Mauro Daniel Rodrigues Paixão, Sebastião Kauã de Sousa Bispo, Natasha Cristina Oliveira Andrade, Juliane Lima Alencar, Tamires de Nazaré Soares, Joyce dos Santos Freitas, Rubenilson Caldas Valois, Ana Lúcia da Silva Ferreira, Karla Valéria Batista Lima, Luana Nepomuceno Gondim Costa Lima

**Affiliations:** Programa de Pós-graduação em Biologia Parasitária na Amazônia, Universidade Estado do Pará e Instituto Evandro Chagas (PPGBPA/UEPA/IEC). Belém, Pará, Brazil; Fundação Santa Casa de Misericórdia do Pará, (FSCMP). Belém, Pará, Brazil; Universidade da Amazônia (UNAMA). Ananindeua, Pará, Brazil; Programa de Pós-graduação em Epidemiologia e Vigilância em Saúde, Instituto Evandro Chagas (PPGEVS/IEC). Ananindeua, Pará, Brazil; Programa de Pós-graduação em Enfermagem, Universidade do Estado do Pará (PPGENF/UEPA). Belém, Pará, Brazil; Faculdade de Enfermagem, Instituto de Ciências de Saúde, Universidade Federal do Pará (FAENF/ICS/UFPA). Belém, Pará Brazil

**Keywords:** Epidemiology, Health Surveillance, Health Indicators, SINASC, Live Births, COVID-19

## Abstract

The surveillance of live births in Brazil has been carried out since 1990 by the Information System on Live Births (SINASC), which was implemented by the Ministry of Health aiming at standardized registration on a national level. The state of Pará is part of the Brazilian Amazon, northern Brazil, which has several unique characteristics. Thus, the purpose of this study was to identify the epidemiological pattern of live births before and during the pandemic of COVID-19 in the state of Pará, 2016 to 2020. This is an ecological epidemiological time-series study, using epidemiological surveillance data from DATASUS, referring to the Live Births Information System (SINASC). These are data that have been treated by surveillance and are in aggregate format. The study population is the live births residing in the state of Pará, in the period from 2016 to 2020. The data collection instrument was the Declaration of Live Births (DLB). There were 689,454 live births, and the highest rates of births were and continued to remain in the Marajó II, Baixo Amazonas, Xingu, and Tapajós regions. The Metropolitan I and Araguaia regions were and continue to be the lowest rates in the state. Age of the mother 15 to 19 years old 22.29%, 20 to 24 years old 30.05% and 25 to 29 years old 22.58%, most of the single pregnancy type 98.32%, prenatal consultations, performed 7 or more 48.10%, followed by 4 to 6 consultations 33.98%, most presented 7 or more years of the study 48.10%, followed by 3 to 6 years 33.98%. Represented 51.21% male and 48.77% female. The occurrence of congenital anomalies represented 0.52% of live births. Another congenital malformation and deformity were the most prevalent at 25.53%, followed by Congenital deformities of the feet 14.90%, Other congenital malformations of the nervous system 14.84%, and Other congenital malformations 10.77%, Cleft lip, and cleft palate 8.88%, Other congenital malformations digestive tract 8.10%. The demographic transition has already occurred for several decades, including the reduction of fertility and birth rate, so our study showed that the reduction in the number of live births was already a reality in the country, but we emphasize that this reduction was enhanced by the pandemic. We observed greater adherence to prenatal care and a lower prevalence of low birth weight compared to other studies, but the limitation was the absence of studies in the same place of the research. Regarding data incompleteness, we emphasize the ignored fields that reflect the fragility in the surveillance of live births, which was reinforced by the literature.

## INTRODUCTION

Surveillance of live births in Brazil has been carried out since 1990 by the Information System on Live Births (SINASC), which was implemented by the Ministry of Health aiming at a standardized national registry of information on live births. SINASC uses the Declaration of Live Births (DLB) as an instrument for data collection, which has several variables on the mother, prenatal care, delivery, and the newborn. This surveillance system represents an essential source of information for health research and evaluation in the maternal and child area [1].

Thus, SINASC surveillance subsidizes public health measures concerning women’s and children’s health, such as public policies to reduce maternal mortality, and adherence to quality prenatal, delivery, and puerperium care. The DLB is mandatory to be issued in three copies, which will be a requirement for the birth certificate, which is a fundamental document for the child’s social, educational, and economic policies regarding society. [2].

Thus, the epidemiological pattern of live births has been studied for several years, because profile changes must be identified for the development of strategies and understanding of associated factors. Authors have discussed the demographic transition, which highlights the reduction in fertility and birth rate, which has been occurring for decades in the world and Brazil [3].

The state of Pará is part of the Brazilian Amazon, northern Brazil, which has several unique characteristics, such as extensive geographical territory, compared to several European countries together, as well as being composed of rural areas larger than urban areas, which hinders access to education, health services, and health surveillance, indigenous peoples, illegal mining, mercury contamination, factors that directly impact the health of the population, making them vulnerable [4–6].

The arrival of the pandemic by COVID-19 weakened health and surveillance services worldwide, and it was no different in Brazil and Pará. However, the state of Pará already has local vulnerabilities, thus the objective of this study was to identify the epidemiological pattern of live births before and during the pandemic of COVID-19 in the state of Pará, 2016 to 2020.

## METHOD

Epidemiological, ecological, and time-series study with data from DATASUS’ epidemiological surveillance of the Live Births Information System (SINASC). These are data that have been treated by surveillance and are in an aggregated format.

The study population is the live births residing in the state of Pará (figure 1), in the period from 2016 to 2020. The data were made available on the website of the Department of Informatics of the Unified Health System (DATASUS) [7].

**Figure 1.**
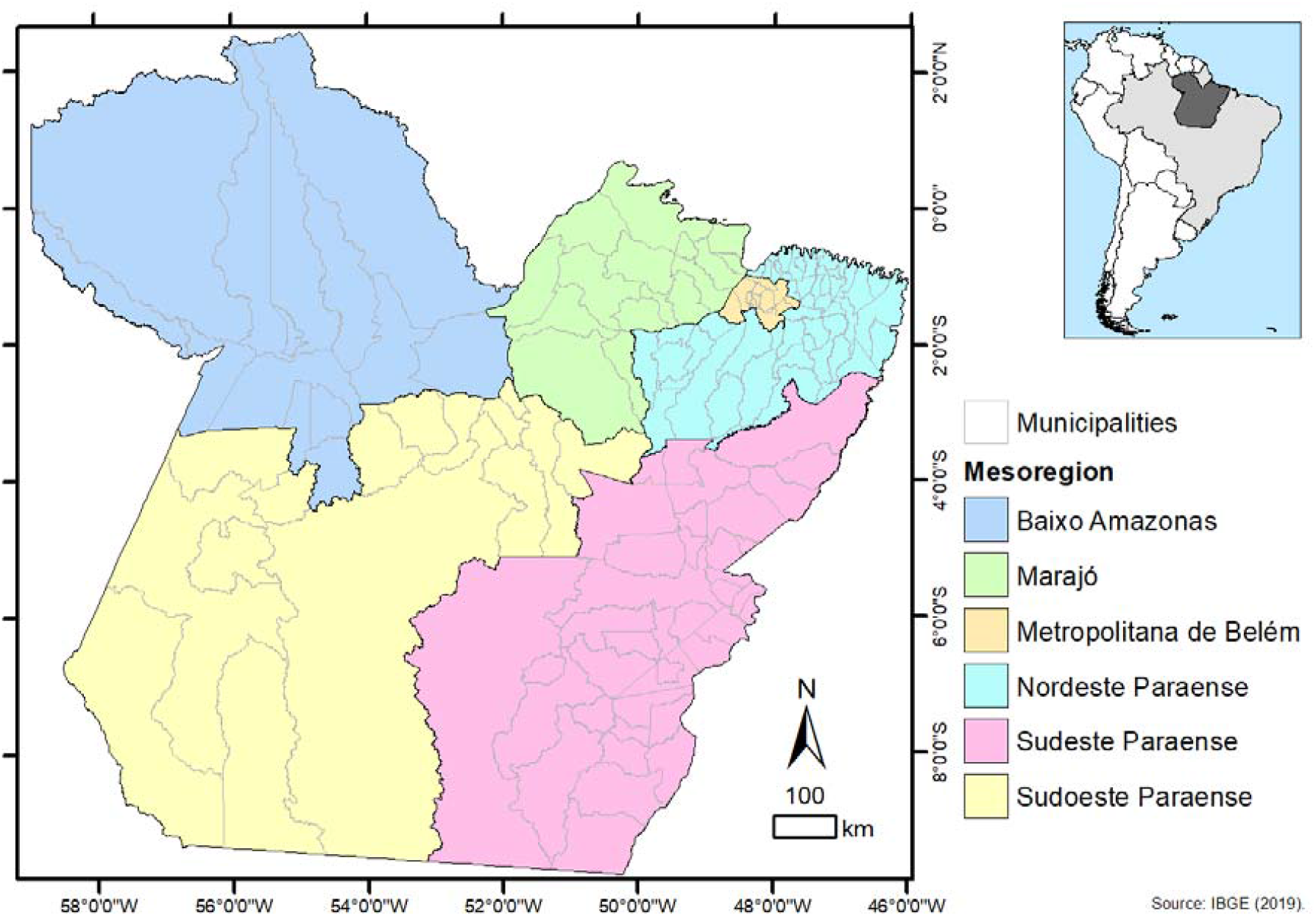
The spatial location of the mesoregions and municipalities of the State of Pará, Amazon, Brazil. Source: (Sardinha, et al. 2021)[8].

The instrument for data collection was the Live Birth Declaration (DNV), the variables, year, age group, type of pregnancy, type of delivery, prenatal consultations, education, congenital anomaly, gender, and type of congenital anomaly were extracted. The data were analyzed by Excel 2019, from absolute and relative numbers, as well as analysis of the curve of the number of live births per year by the R2 equation, which shows if there is a change in pattern and informs the percentage of the difference between the years. We performed the calculation of the birth rate by health region from the resident population also extracted from DATASUS, the calculation was:

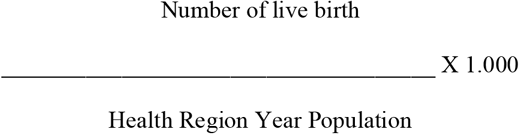

The spatial distribution of live births was performed by the health region of the state of Pará in the ArcGIS software (https://www.arcgis.com/) and classified according to the results of the birth rates, in five classes in red.

According to Resolution No. 510 of April 7, 2016, Article II, which deals with research that uses publicly accessible data, under Law No. 12,527 of November 18, 2011, Articles III (research that uses information in the public domain) and V (research in databases whose information is aggregated, without the possibility of individual identification), will not be registered or evaluated by the Ethics and Research Committee (CEP/CONEP) system. Thus, these types of studies are not recommended to be submitted for ethical review and can be freely conducted, since the publicly available data does not include data such as the names, phone numbers, and addresses of the participants [9,10].

## RESULTS

In the state of Pará, there were 689,454 live births in the study period, highlighting the drop in the year 2020 which was 132,937, compared to 2019 that where 138,338. The health regions with the highest numbers were Metropolitan I with 21.32%, Carajás with 11.26%, and Baixo Amazonas with 11.22%, proportional to being the most populous regions (table 1).

**Table 1.** Number of live births in the state of Pará by health region, from 2016 to 2020.

| Health Region | 2016 | 2017 | 2018 | 2019 | 2020 | Total | % |
| --- | --- | --- | --- | --- | --- | --- | --- |
| Araguaia | 7477 | 7812 | 8311 | 8020 | 7731 | 39351 | 5,71 |
| Baixo Amazonas | 14998 | 15422 | 15697 | 15810 | 15456 | 77383 | 11,22 |
| Carajás | 15629 | 15330 | 16130 | 15335 | 15224 | 77648 | 11,26 |
| Lago de Tucuruí | 6915 | 6844 | 6718 | 6474 | 6282 | 33233 | 4,82 |
| Metropolitana I | 30359 | 30529 | 30197 | 29137 | 26753 | 146975 | 21,32 |
| Metropolitana II | 6136 | 5981 | 6216 | 6106 | 5725 | 30164 | 4,38 |
| Metropolitana III | 13650 | 13605 | 14186 | 14109 | 13264 | 68814 | 9,98 |
| Rio Caetés | 8716 | 8557 | 8587 | 8314 | 8093 | 42267 | 6,13 |
| Tapajós | 3944 | 4409 | 4381 | 4360 | 4491 | 21585 | 3,13 |
| Tocantins | 11523 | 11768 | 12227 | 11957 | 11605 | 59080 | 8,57 |
| Xingu | 7009 | 6795 | 6970 | 6836 | 6575 | 34185 | 4,96 |
| Marajó I | 3969 | 3959 | 4075 | 4038 | 4025 | 20066 | 2,91 |
| Marajó II | 7354 | 7671 | 8123 | 7842 | 7713 | 38703 | 5,61 |
| Total | 137679 | 138682 | 141818 | 138338 | 132937 | 689454 | 100,00 |

Source: MS/SVS/DASIS - Live Births Information System - SINASC.

In the analysis by the number of live births per year, the trend line showed the reduction of live births each year, enhanced by 2020. The R2 value showed that each year the reduction tends to be 23% (graph 1).

**Graph 1.**
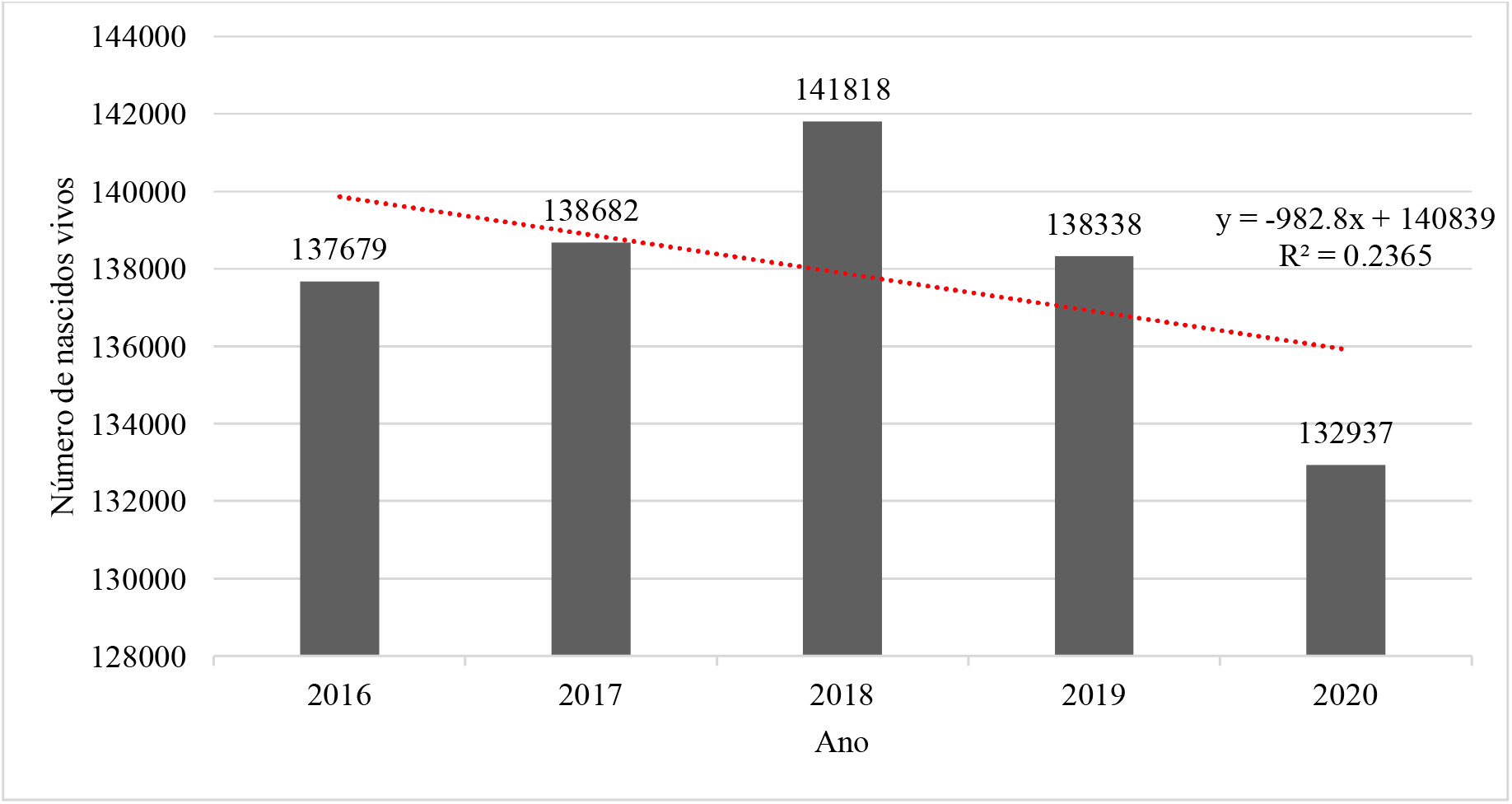
Number of live births per year, in the state of Pará from 2016 to 2020. Source: MS/SVS/DASIS - Live Births Information System - SINASC.

We calculated the birth rate by health region, and also presented the spatial distribution. The highest birth rates were and remain in the regions Marajó II (per year 23.86/24.58/25.69/24.49/23.80), Baixo Amazonas (per year 19.87/20.28/20.49/20.49/19.89), Xingu (per year 20.67/19.82/20.11/19.52/18.58 and Tapajós (per year 18.01/20.07/19.87/19.72/20.25). The Metropolitan I (per year 13.85/13.83/13.58/13.02/11.87) and Araguaia (per year 13.84/14.23/14.90/14.15/13.44) regions were and still are the lowest rates in the state of Pará (table 2) (graph 2) (figures 1,2 and 3).

**Graph 2.**
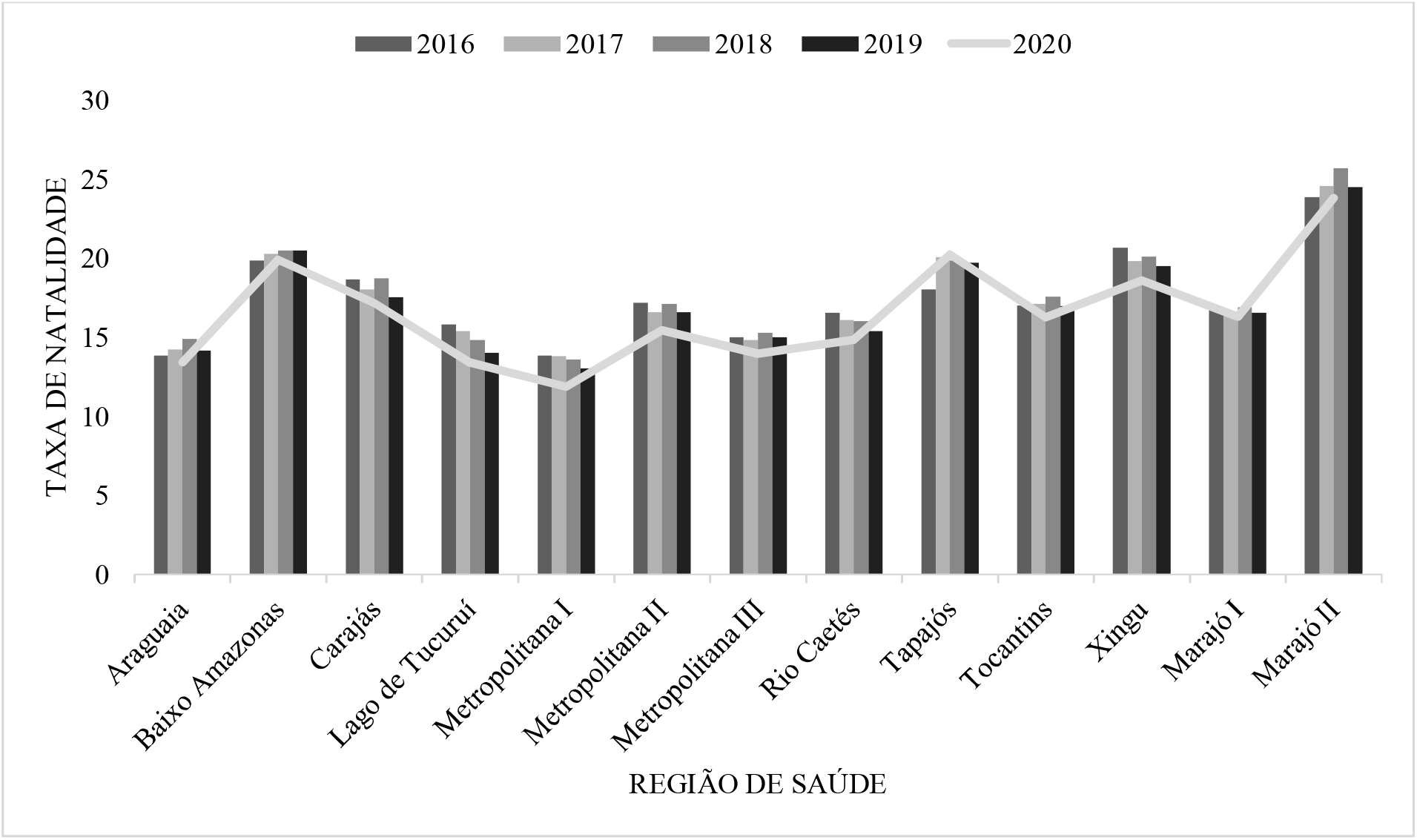
Birth Rate per 1,000 inhabitants, health region in the state of Para 2016 to 2020. Source: MS/SVS/DASIS - Live Births Information System - SINASC.

**Figure 1.**
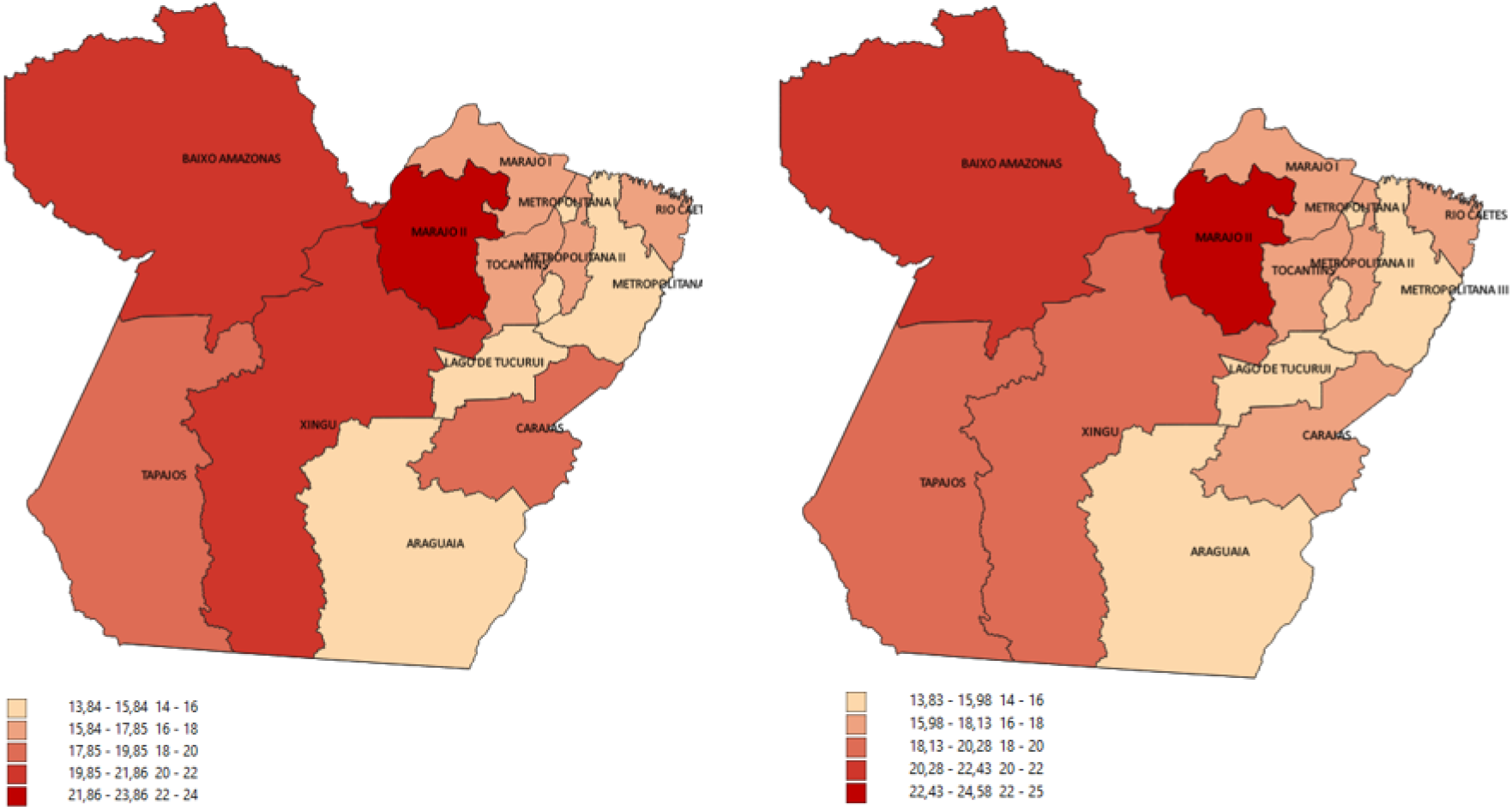
Spatial distribution of birth rate by health region in the state of Pará, 2016 and 2017. Source: MS/SVS/DASIS - Live Births Information System - SINASC. Software ArcGIS (https://www.arcgis.com/).

**Figure 2.**
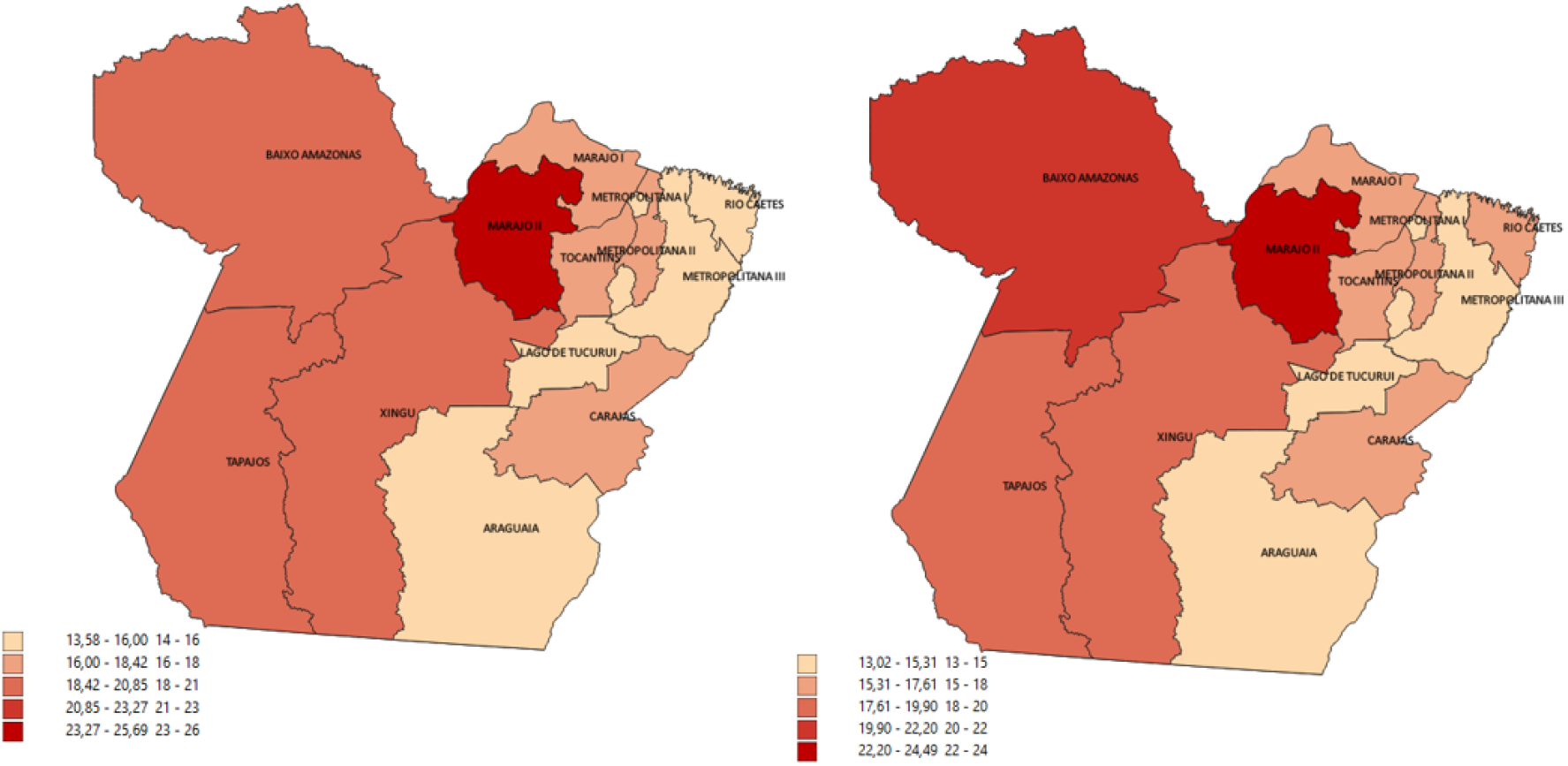
Spatial distribution of birth rate by health region in the state of Pará, 2018 and 2019. Source: MS/SVS/DASIS - Live Births Information System - SINASC. Software ArcGIS https://www.arcgis.com/).

**Figure 3.**
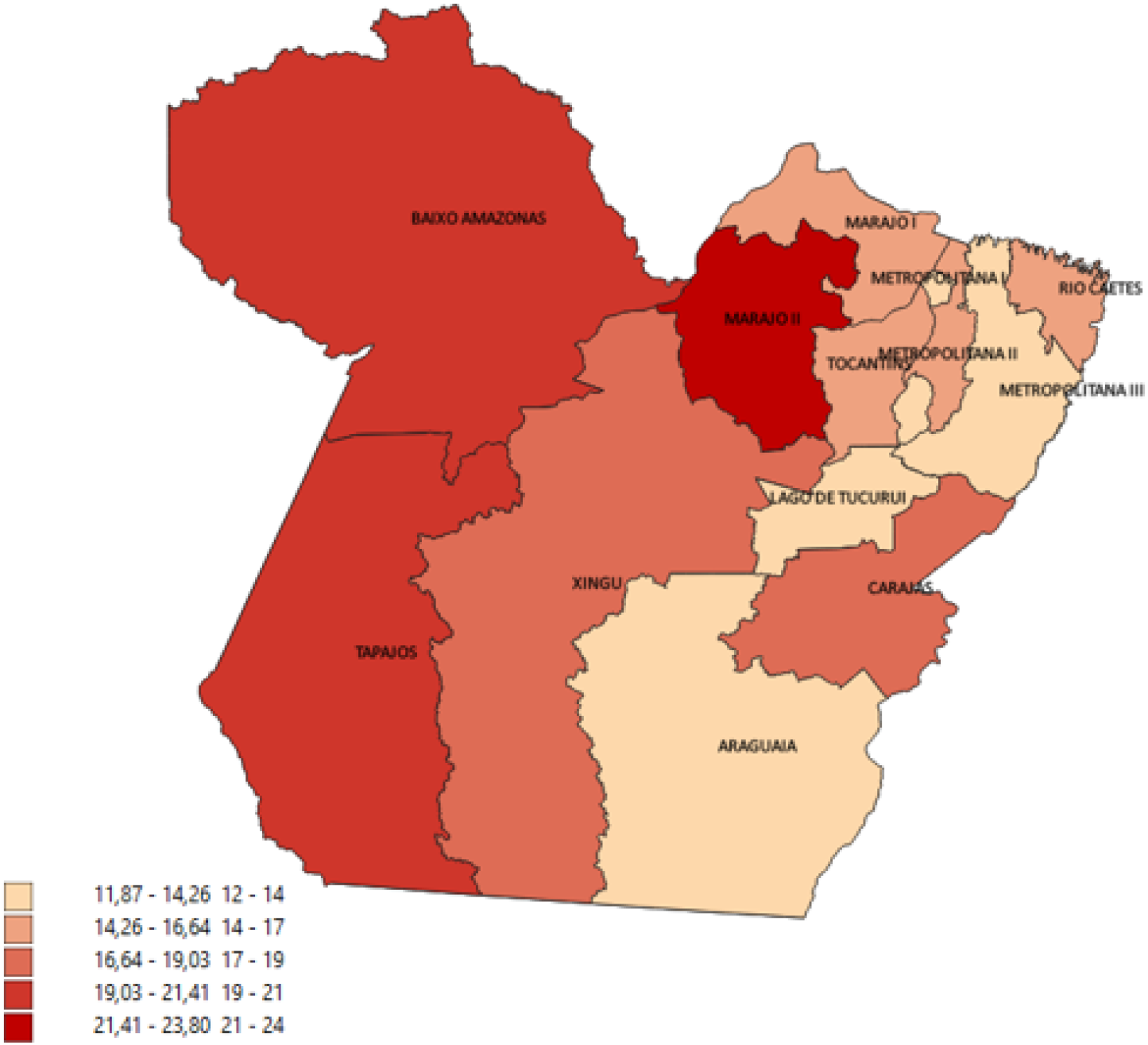
Spatial distribution of birth rate by health region in the state of Pará, 2020. Source: MS/SVS/DASIS - Live Births Information System - SINASC. Software ArcGIS (https://www.arcgis.com/).

**Table 2.** Birth Rate per 1,000 population, health region in the state of Para 2016 to 2020.

| Health Region | 2016 | 2017 | 2018 | 2019 | 2020 |
| --- | --- | --- | --- | --- | --- |
| Araguaia | 13.84 | 14.23 | 14.90 | 14.15 | 13.44 |
| Baixo Amazonas | 19.87 | 20.28 | 20.49 | 20.49 | 19.89 |
| Carajás | 18.66 | 18.03 | 18.70 | 17.52 | 17.15 |
| Lago de Tucuruí | 15.82 | 15.37 | 14.81 | 14.03 | 13.38 |
| Metropolitana I | 13.85 | 13.83 | 13.58 | 13.02 | 11.87 |
| Metropolitana II | 17.19 | 16.59 | 17.08 | 16.61 | 15.43 |
| Metropolitana III | 15.02 | 14.81 | 15.27 | 15.02 | 13.97 |
| Rio Caetés | 16.53 | 16.09 | 16.00 | 15.36 | 14.83 |
| Tapajós | 18.01 | 20.07 | 19.87 | 19.72 | 20.25 |
| Tocantins | 16.98 | 17.12 | 17.56 | 16.96 | 16.26 |
| **Xingu** | 20.67 | 19.82 | 20.11 | 19.52 | 18.58 |
| **Marajó I** | 16.88 | 16.63 | 16.91 | 16.55 | 16.30 |
| **Marajó II** | 23.86 | 24.58 | 25.69 | 24.49 | 23.80 |
| **Total** | 16.52 | 16.47 | 16.66 | 16.08 | 15.30 |

Source: MS/SVS/DASIS - Live Births Information System - SINASC.

Regarding sex, the live births in the study period represented (51.21%) male and (48.77%) female, the remaining were ignored in the sex field. Regarding the age range, the majorities of women were between 15 to 19 years (22.29%), 20 and 24 years (30.05%), and 25 to 29 years (22.58%). As well as the majority of single pregnancy type 98.32%. The mother’s schooling by the number of years of study, the majority presented 7 or more years (48.10%), followed by 3 to 6 years (33.98%). In the comparison of schooling with the number of prenatal visits, 8 to 11 years of study who made 7 visits or more (59.19%), and those who did not make any visits with the same schooling was (35.78%) (table 3).

**Table 3.**
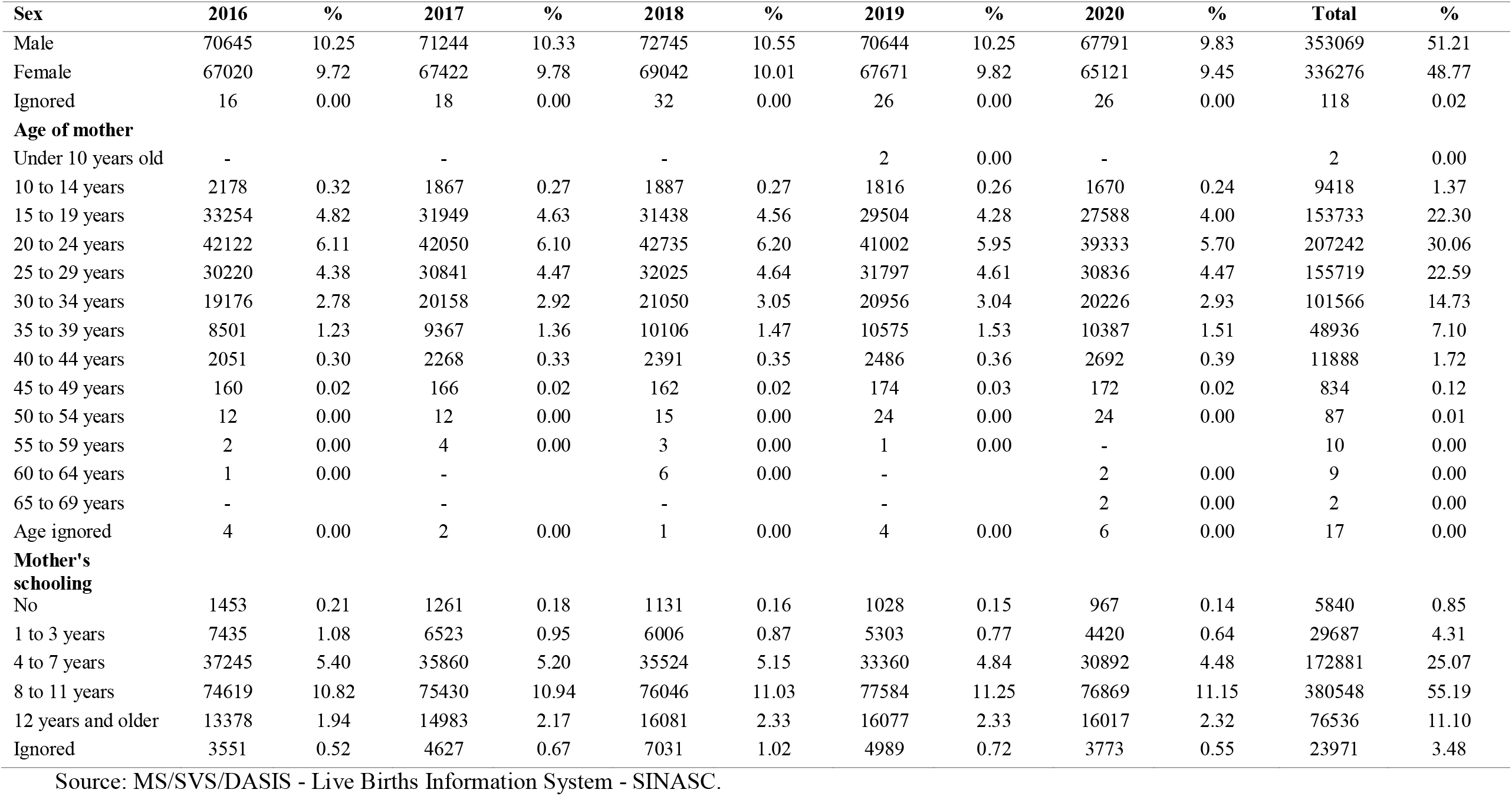
Sociodemographic variables of the mother and births by health region, state of Pará, 2016 to 2020.

The single pregnancy type accounted for 98.33% of the live births. About the type of delivery, 50.20% were vaginal and 49.70 were cesarean. Concerning prenatal consultations, the majority had 7 or more 48.10%, followed by 4 to 6 consultations 33.98%. Regarding birth weight, the study showed 52,378 (7.59%) live births with a low birth weight of <2,499 g. The majority 64.61% weigh between 3000 to 3999 g (table 4).

**Table 4.** Pregnancy and Birth Variables by health region, state of Pará, 2016 to 2020.

| **Pregnancy Type** | **2016** | **%** | **2017** | **%** | **2018** | **%** | **2019** | **%** | **2020** | **%** | **Total** | **%** |
| --- | --- | --- | --- | --- | --- | --- | --- | --- | --- | --- | --- | --- |
| Single | 135392 | 19.64 | 136509 | 19.80 | 139408 | 20.22 | 135921 | 19.71 | 130702 | 18.96 | 677932 | 98.33 |
| Double | 2058 | 0.30 | 1969 | 0.29 | 2202 | 0.32 | 2254 | 0.33 | 2071 | 0.30 | 10554 | 1.53 |
| Triple and more | 16 | 0.00 | 26 | 0.00 | 39 | 0.01 | 35 | 0.01 | 33 | 0.00 | 149 | 0.02 |
| Ignored | 215 | 0.03 | 180 | 0.03 | 170 | 0.02 | 131 | 0.02 | 132 | 0.02 | 828 | 0.12 |
| **Type of birth** |  |  |  |  |  |  |  |  |  |  |  |  |
| Vaginal | 71213 | 10.33 | 70954 | 10.29 | 71553 | 10.38 | 68702 | 9.96 | 63701 | 9.24 | 346123 | 50.20 |
| Cesarean | 66259 | 9.61 | 67573 | 9.80 | 70139 | 10.17 | 69535 | 10.09 | 69144 | 10.03 | 342650 | 49.70 |
| Ignored | 209 | 0.03 | 157 | 0.02 | 127 | 0.02 | 104 | 0.02 | 93 | 0.01 | 690 | 0.10 |
| **Prenatal consultation** |  |  |  |  |  |  |  |  |  |  |  |  |
| None | 8241 | 1.20 | 6586 | 0.96 | 6401 | 0.93 | 5457 | 0.79 | 7653 | 1.11 | 34338 | 4.98 |
| From 1 to 3 | 16809 | 2.44 | 17580 | 2.55 | 17428 | 2.53 | 15836 | 2.30 | 18649 | 2.70 | 86302 | 12.52 |
| From 4 to 6 | 48595 | 7.05 | 48056 | 6.97 | 48342 | 7.01 | 45203 | 6.56 | 44095 | 6.40 | 234291 | 33.98 |
| 7 or more | 63519 | 9.21 | 65987 | 9.57 | 69028 | 10.01 | 71451 | 10.36 | 61660 | 8.94 | 331645 | 48.10 |
| Ignored | 517 | 0.07 | 475 | 0.07 | 620 | 0.09 | 394 | 0.06 | 881 | 0.13 | 2887 | 0.42 |
| **Birth Weight** |  |  |  |  |  |  |  |  |  |  |  |  |
| Less than 500g | 208 | 0.03 | 198 | 0.03 | 225 | 0.03 | 221 | 0.03 | 200 | 0.03 | 1052 | 0.15 |
| 500 to 999g | 481 | 0.07 | 553 | 0.08 | 546 | 0.08 | 556 | 0.08 | 507 | 0.07 | 2643 | 0.38 |
| 1000 to 1499g | 846 | 0.12 | 884 | 0.13 | 901 | 0.13 | 837 | 0.12 | 832 | 0.12 | 4300 | 0.62 |
| 1500 a 2499g | 8687 | 1.26 | 8868 | 1.29 | 9180 | 1.33 | 8982 | 1.30 | 8666 | 1.26 | 44383 | 6.44 |
| 2500 a 2999g | 30810 | 4.47 | 30259 | 4.39 | 31165 | 4.52 | 30285 | 4.39 | 29129 | 4.22 | 151648 | 22.00 |
| 3000 to 3999 g | 88833 | 12.88 | 89820 | 13.03 | 91449 | 13.26 | 89489 | 12.98 | 85902 | 12.46 | 445493 | 64.61 |
| 4000g and above | 7664 | 1.11 | 7942 | 1.15 | 8197 | 1.19 | 7871 | 1.14 | 7579 | 1.10 | 39253 | 5.69 |
| Ignored | 152 | 0.02 | 160 | 0.02 | 156 | 0.02 | 100 | 0.01 | 123 | 0.02 | 691 | 0.10 |

Source: MS/SVS/DASIS - Live Births Information System - SINASC.

The occurrence of congenital anomalies represented 0.52% of live births, 0.55% male, and 0.47% female (table 6). We analyzed the types of congenital anomalies, Other congenital malformation and deformity were the most prevalent 25.53%, followed by Congenital deformities of the feet 14.90%, Other congenital malformations of the nervous system 14.84%, Other congenital malformations 10.77%, Cleft lip and cleft palate 8.88%, Other congenital malformations digestive tract 8.10%. The others represent less than 6% each (table 5 and 6).

**Table 5.** Occurrence of congenital anomalies by health region, state of Pará, 2016 to 2020.

| **Congenital anomaly** | **2016** | **%** | **2017** | **%** | **2018** | **%** | **2019** | **%** | **2020** | **%** | **Total** | **%** |
| --- | --- | --- | --- | --- | --- | --- | --- | --- | --- | --- | --- | --- |
| Yes | 626 | 0.09 | 699 | 0.10 | 778 | 0.11 | 753 | 0.11 | 757 | 0.11 | 3613 | 0.52 |
| No | 135516 | 19.66 | 137475 | 19.94 | 140105 | 20.32 | 135385 | 19.64 | 129587 | 18.80 | 678068 | 98.35 |
| Ignored | 1539 | 0.22 | 510 | 0.07 | 936 | 0.14 | 2203 | 0.32 | 2594 | 0.38 | 7782 | 1.13 |
| **Type of congenital anomaly** |  |  |  |  |  |  |  |  |  |  |  |  |
| Spina bifida | 16 | 0.00 | 20 | 0.00 | 27 | 0.00 | 26 | 0.00 | 20 | 0.00 | 109 | 0.02 |
| Other congenital malformations of the nervous system | 126 | 0.02 | 109 | 0.02 | 122 | 0.02 | 95 | 0.01 | 83 | 0.01 | 535 | 0.08 |
| Congenital malformations of the circulatory system | 9 | 0.00 | 17 | 0.00 | 29 | 0.00 | 40 | 0.01 | 43 | 0.01 | 138 | 0.02 |
| Cleft lip and cleft palate | 59 | 0.01 | 68 | 0.01 | 66 | 0.01 | 59 | 0.01 | 68 | 0.01 | 320 | 0.05 |
| Absent atresia and stenosis of the small intestine | - |  | 6 | 0.00 | 3 | 0.00 | 2 | 0.00 | 2 | 0.00 | 13 | 0.00 |
| Other congenital malformations of the digestive system | 30 | 0.00 | 43 | 0.01 | 63 | 0.01 | 79 | 0.01 | 77 | 0.01 | 292 | 0.04 |
| Undescended testicle | 3 | 0.00 | 4 | 0.00 | 4 | 0.00 | 7 | 0.00 | 7 | 0.00 | 25 | 0.00 |
| Other genitourinary system malformations | 29 | 0.00 | 29 | 0.00 | 48 | 0.01 | 42 | 0.01 | 39 | 0.01 | 187 | 0.03 |
| Congenital deformities of the hip | - |  | - |  | 1 | 0.00 | 4 | 0.00 | - |  | 5 | 0.00 |
| Congenital deformities of the feet | 95 | 0.01 | 102 | 0.01 | 115 | 0.02 | 118 | 0.02 | 107 | 0.02 | 537 | 0.08 |
| Other congenital malformations and deformities of the musculoskeletal system | 164 | 0.02 | 201 | 0.03 | 189 | 0.03 | 171 | 0.02 | 195 | 0.03 | 920 | 0.13 |
| Other congenital malformations | 77 | 0.01 | 67 | 0.01 | 76 | 0.01 | 80 | 0.01 | 88 | 0.01 | 388 | 0.06 |
| Chromosomal anomalies NCOP | 15 | 0.00 | 30 | 0.00 | 30 | 0.00 | 27 | 0.00 | 27 | 0.00 | 129 | 0.02 |
| Hemangioma and lymphangioma | - |  | 1 | 0.00 | 3 | 0.00 | 2 | 0.00 | - |  | 6 | 0.00 |
| No congenital anomaly/not informed | 137058 | 19.88 | 137987 | 20.01 | 141043 | 20.46 | 137589 | 19.96 | 132182 | 19.17 | 685859 | 99.48 |

Fonte: MS/SVS/DASIS - Sistema de Informações sobre Nascidos Vivos – SINASC.

**Table 6.** Live births by type of congenital anomaly, Pará state 2016 to 2020.

| **Congenital anomaly type** | **Total**<br>**(3604)** | **%** |
| --- | --- | --- |
| **Other congenital malformations and deformities of the musculoskeletal system** | 920 | 25.53 |
| **Congenital deformities of the feet** | 537 | 14.9 |
| **Other congenital malformations of the nervous system** | 535 | 14.84 |
| **Other congenital malformations** | 388 | 10.77 |
| **Cleft lip and cleft palate** | 320 | 8.88 |
| **Other congenital malformations of the digestive tract** | 292 | 8.1 |
| **Other congenital malformations of the genitourinary system** | 187 | 5.19 |
| **Congenital malformations of the circulatory system** | 138 | 3.83 |
| **Chromosomal abnormalities NCOP** | 129 | 3.58 |
| **Spina bifida** | 109 | 3.02 |
| **Undescended testicle** | 25 | 0.69 |
| **Absent atresia and stenosis of the small bowel** | 13 | 0.36 |
| **Hemangioma and lymphangioma** | 6 | 0.17 |
| **Congenital hip deformities** | 5 | 0.14 |

Source: MS/SVS/DASIS - Live Births Information System - SINASC.

## DISCUSSION

In this study, we characterized the live births of the state of Pará in the years 2016 to 2020, based on data from the epidemiological surveillance of live births in Brazil SINASC. On the number of live births, we evidenced a reduction in the trend line, which was potentiated in 2020. According to the civil registry, an official page that shares data on births shows that in 2021 in the state of Pará there were 121,739 births, and highlights that with the arrival of the pandemic in Brazil births reduced by 15%, but emphasizes that the birth rate decline had already been occurring [11].

A report in the CNN Brazil newspaper emphasized the reduction of the fertility rate in Brazil, which was enhanced by the pandemic of COVID-19. They highlighted that the fear and anguish of staying in a maternity ward in times of pandemic, emotional issues, and worries about SARS-CoV-2 are associated with this reduction [12].

Studies also showed that maternal mortality increased during the pandemic and that it was directly associated with COVID-19. In 2021, Brazil had the highest maternal mortality rate for COVID-19 in the world, which alerted researchers, because due to underreporting these data can be even higher, as COVID-19 is potentiated during pregnancy due to various physiological changes [13,14]. Research on maternal mortality in the state of Pará showed that 44 maternal deaths had occurred by June 8, 2020, of which 20 were due to COVID-19. [15].

Regarding the birth rate reduction, a 2012 study already discussed the demographic transition since 1950, highlighting the declines in mortality, birth rate, and fertility in this process, but emphasized that in the North and Northeast of Brazil this process was less evident [16]. Another study analyzed the fertility transition based on epidemiological data, and stated that the drop in fertility began in the mid-1930s in Brazil, especially in the South and Southeast regions, and also cited in the study factors regarding the reduction, such as women’s choice to have fewer children, low social cost, contraceptive methods, and education [17].

Research in the municipality of Rio de Janeiro on the epidemiological profile of live births showed that most were full-term, adequate weight, and Apgar score on the first and fifth minutes between 7 and 10 points. The mothers were mostly brown, single, with 8 to 11 years of schooling, and aged between 20 and 34. Regarding the type of delivery, Cesarean was more frequent [18]. Being similar concerning age, and education, however different in the type of delivery, because in our study the predominance was a vaginal delivery, even with little difference between the cesarean delivery. Another study described the profile of live births in the city of Viçosa from 2001 to 2007, showing the main age range of mothers was between 20 and 29 years, representing 55% of this age group in 2007. The percentage of teenage mothers decreased from 18.2 to 14.6% from 2001 to 2007. Concerning the type of delivery, an increase in cesarean sections and a decrease in vaginal deliveries are emphasized. However, vaginal deliveries are higher than cesarean deliveries among adolescents. As for weight, low birth weight reached 8% in 2007. The Apgar score in the first and fifth minutes >7 represented 73 and 89% in 2007. [19].

A study aimed to know the epidemiological profile of births in Chapecó/SC, in the period from July 2011 to June 2013, based on SINASC. They identified a total of 5,918 live births, of these 9.2% were born weighing less than 2,500 g; 9.4% were premature; 15.7% were children of adolescent mothers, and 20% of women had seven prenatal visits or less [20]. In our study, the majority had 7 or more prenatal visits, and the prevalence of low weight was lower.

Regarding the prevalence of congenital anomalies, a study described congenital anomalies (CA) among live births of mothers residing in Tangará da Serra, MT, Brazil, the period 2006-2016. Of 15,689 live births, 77 were registered (prevalence of 4.9/1,000); there was an 80.7% increase in CA registered in 2016, representing 10.3/1. 000 live births, including five cases of microcephaly; The prevalence of CA was higher among children born to women aged over 35 years (prevalence ratio [PR] =1.91; confidence interval [95% CI] 1.01;3.60), premature infants (PR=2.22; 95% CI 1.26;3.92) and low birth weight infants (PR=3.21; 95% CI 1.86;5.54) [21].

Concerning the higher birth rates in the health regions of Marajó II, Baixo Amazonas, and the Tapajós, they are associated with lower schooling and less access to health services that these regions have characteristics of local vulnerability because they have a large geographical extension and have many people in rural and indigenous areas. It is worth mentioning that these are regions of illegal mining and environmental contamination from mercury [22,23].

Regarding the quality of the SINASC information, a study highlighted the weaknesses in the completeness of the essential fields in the surveillance of live births, and that these indicators are worse in the northern region of the country [24]. In another study in Brazil from 2006 to 2010 on the completeness of the variables of the DNV, 21 of the 23 variables analyzed showed completeness above 90.0%. 97.9% of the hospital delivery variables had complete data; they found no differences in the proportion of births concerning macroregion and sex concerning the 2010 census; 82.6% of the data were received on time in 2010; the ratio between live births notified and estimated was 89.4% in 2006 and 97.4% in 2010 [25]. Another study conducted a review of data completeness, from previously published studies, 13 articles were reviewed. The evaluation of coverage was the subject of analysis in eight studies, completeness in four, and reliability in seven. Most of them presented results of coverage higher than 90%, indicating their feasibility for the calculation of indicators. However, under-registration of births in SINASC prevailed, ranging from 75.8% to 99.5%. The variables maternal education, parity, and the number of prenatal visits were those that presented the greatest inconsistency. Thus, the variable parity was the one that presented the greatest incompleteness [26].

The impossibility of including the data of live births from 2021 stands out as a limitation because the DATASUS surveillance system only makes the data available after the treatment of the data and the qualification of the information, and in Brazil generally the availability of data from the previous year is only made in October to December of the current year.

## CONCLUSION

We presented the epidemiological pattern of live births in the state of Pará, before and during the pandemic of COVID-19, and showed that the demographic transition had already been occurring for several decades, including the reduction of fecundity and birth rate, thus our study showed that the reduction in the number of live births was already a reality in the country, but we emphasized that this reduction was enhanced by the pandemic.

The other variables were similar, except for the higher prenatal care adherence which was better than the previous studies, and the lower prevalence of low weight, but the limitation was that studies in the same place of the research were not found.

Regarding the incompleteness of the data, we highlight the ignored fields that reflect the fragility in the surveillance of live births, which was reinforced by the literature. Health promotion should be strengthened concerning the qualification of surveillance professionals, and the theme should be discussed with greater accuracy in the academies for training health professionals.

## Data Availability

DATASUS

https://datasus.saude.gov.br/

## Notes

### Competing Interest Statement

The authors have declared no competing interest.

### Funding Statement

This study was funded by Conselho Nacional de Desenvolvimento Cientifico e Tecnologico (CNPQ), Coordenacao de Aperfecoamento de Pessoal de Nivel Superior (CAPES) and Fundacao Amazonia de Amparo a Estudos e Pesquisas (FAPESPA)

### Author Declarations

According to Resolution No. 510 of April 7, 2016, Article II, which deals with research that uses publicly accessible data, under Law No. 12,527 of November 18, 2011, Articles III (research that uses information in the public domain) and V (research in databases whose information is aggregated, without the possibility of individual identification), will not be registered or evaluated by the Ethics and Research Committee system. Thus, these types of studies are not recommended to be submitted for ethical review and can be freely conducted, since the publicly available data does not include data such as the names, phone numbers, and addresses of the participants

